# Surveillance of acute SARS-CoV-2 infections in school children and point-prevalence during a time of high community transmission in Switzerland

**DOI:** 10.1101/2020.12.24.20248558

**Authors:** Susi Kriemler, Agne Ulyte, Priska Ammann, Gabriela P. Peralta, Christoph Berger, Milo A Puhan, Thomas Radtke

## Abstract

**Background:** Switzerland had one of the highest incidence of severe acute respiratory syndrome coronavirus 2 (SARS-CoV-2) infections in Europe during the second wave. Schools were open as in most of Europe with specific preventive measures in place. However, the frequency and transmission of acute unrecognized, asymptomatic or oligosymptomatic infections in schools during this time of high community transmission is unknown. Thereof, our aim was to pilot a surveillance system that detects acute SARS-CoV-2 infections in schools and possible transmission within classes.

**Methods:** 14 out of the randomly selected sample of the Ciao Corona cohort study participated between December 1 and 11, a time when incidence rate for SARS-CoV-2 infections was high for the canton of Zurich. We determined point-prevalence of acute SARS-CoV-2 infections of school children attending primary and secondary school. A buccal swab for polymerase chain reaction (PCR) and a rapid diagnostic test (RDT) to detect SARS-CoV-2 were taken twice 1 week apart (T1 and T2) in a cohort of children from randomly selected classes. A questionnaire assessed demographics and symptoms compatible with a SARS-CoV-2 infection during the past 5 days.

**Results:** Out of 1299 invited children, 641 (49%) 6- to 16-year-old children and 66 teachers from 14 schools and 67 classes participated in at least 1 of 2 testings. None of the teachers but 1 child had a positive PCR at T1, corresponding to a point-prevalence in children of 0.2% (95% CI 0.0% to 1.1%), and no positive PCR was detected at T2. The child with positive PCR at T1 was negative on the RDT at T1 and both tests were negative at T2. There were 7 (0.6%) false positive RDTs in children and 2 (1.7%) false positive RDTs in teachers at T1 or T2 among 5 schools (overall prevalence 0.7%). All 9 initially positive RDTs were negative in a new buccal sample taken 2 hours to 2 days later, also confirmed by PCR. 35% of children and 8% of teachers reported mild symptoms during the 5 days prior to testing.

**Conclusion:** In a setting of high incidence of SARS-CoV-2 infections, unrecognized virus spread within schools was very low. Schools appear to be safe with the protective measures in place (e.g., clearly symptomatic children have to stay at home, prompt contact tracing with individual and class-level quarantine, and structured infection prevention measures in school). Specificity of the RDT was within the lower boundary of performance and needs further evaluation for its use in schools. Given the low point prevalence even in a setting of very high incidence, a targeted test, track, isolate and quarantine (TTIQ) strategy for symptomatic children and school personnel adapted to school settings is likely more suitable approach than surveillance on entire classes and schools.

**Trial registration:** ClinicalTrials.gov NCT04448717 https://clinicaltrials.gov/ct2/show/NCT04448717

## INTRODUCTION

The role of schools in severe acute respiratory syndrome coronavirus-2 (SARS-CoV-2) infection and transmission is highly debated and ranges from “there is no evidence of danger” [1] to “overlooked or at risk” [2, 3]. This pronounced contradiction has its roots in a puzzle that has many missing pieces. Children are clinically much less severely affected than adults if infected by SARS-CoV-2; 1% of those infected are hospitalized, 0.1% need intensive care and <0.1% die [4]. An asymptomatic or oligosymptomatic infection is present in at least half of the children [5] which allows them to attend school despite undiscovered acute SARS-CoV-2 infection. As asymptomatic cases are so common among children, subtle undiscovered spread may occur, especially in schools where groups of children gather, interact and spend most of their daytime.

Population-based studies on acute infection and seroprevalence show that rates of infection with SARS-CoV-2 in children are similar to those in adults [6-8]. School outbreaks have been described [2], but it is not clear how frequently they occur and how much they contribute to the spread of SARS-CoV-2 on a population level. Retrospective modelling studies based on many assumptions are highly controversial [1, 3, 9, 10]. Some groups have shown that closing schools and universities without making a distinction between these different settings, was associated with a 38% reduction of the reproduction factor R during the first wave of SARS-CoV-2 pandemic [3]. Others documented that school closures alone prevented only 2-4% of deaths [1]. Irrespective of outcomes on morbidity and mortality rates on the population level, school closures come with an enormous burden of social, educational and psychological constraints to millions of children worldwide, and especially to those that need the school system most to be carried and supported by society [11]. Other less disruptive, potentially effective approaches of infection prevention measures can be taken up by schools before a hard lock-down is considered. For example in Switzerland, parents are asked to keep their children at home when sick, each school needs to have protective measures in place to prevent transmissions and specific regulations of authorities are in place. Unfortunately, their impact has been poorly studied [12, 13].

Surveillance of infections in school children may help to better understand the situation in schools, evaluate and adjust protective measures and trigger test, track, isolate and quarantine (TTIQ). Some surveillance studies used antibody tests to determine seroprevalence of SARS-CoV-2 in children [14-16] with few of these studies specifically focusing on the school setting [17]. We recently reported about the protocol and results of the large, population and school-based study Ciao Corona [8, 18, 19]. These epidemiological, serological studies are very valuable to determine seroprevalence and its regional variability, predictors of seropositivity as well as the extent to which clustering of cases occurs within classes and schools over time. But several additional questions cannot be addressed. For example, the number of children in schools with acute unrecognized, asymptomatic SARS-CoV-2 infections and to what extent these children transmit the virus in the school setting is unclear and can only be elucidated using viral testing as done in several outbreak case studies but very few school-based studies [20, 21]. Also, it is unclear how surveillance of such acute but almost silent infections can be linked to TTIQ in order to breaking transmission chains.

Surveillance for acute SARS-CoV-2 infections in school settings can be implemented using polymerase chain reaction (PCR) or rapid diagnostic tests (RDT) [22]. PCR has the advantage of being highly sensitive and detect infections in asymptomatic persons. On the other side, PCR tests take time to deliver results. Potentially infectious school children may transmit the virus until the result is available since sending them into quarantine until the test results are available is not an option in a surveillance setting like for symptomatic persons. Also, a substantial proportion of infected persons still show positive PCR tests while not being infectious anymore. This carries the risk of unnecessary TTIQ [23]. This latter potential problem of PCR tests may be smaller for RDT because they are less sensitive [22] and would allow triggering immediate TTIQ when the test is positive.

The aim of this study was threefold. Firstly, to determine the point prevalence of asymptomatic or oligosymptomatic acute SARS-CoV-2 infections in school children during a time of high community transmission of the SARS-CoV-2. Secondly, to evaluate the feasibility of a surveillance system to monitor acute SARS-CoV-2 infections and clustering in schools. Thirdly, to find out whether a RDT can be used in school in children and their classroom teachers that are asymptomatic or mildly symptomatic to differentiate between the infectious and postinfectious PCR-positive state and how PCR and RDT may be linked to TTIQ in school settings.

## MATERIALS AND METHODS

This observational prospective study was nested in the Ciao Corona cohort study and took part from December 1 to December 11, 2020 at a time of high community transmission of the second wave of the SARS-CoV-2 pandemic. Switzerland is one of the most affected countries worldwide with 4-5000 per 100’000 new cases per day, 300 per 100’000 in the age range 10 to 19 years [24]. The study protocol and related documents were approved by the cantonal ethical committee of Canton Zurich (BASEC Nr. 2020-01336). All parents and teachers gave written and children verbal consent to participate. We invited 15 out of 55 schools from the Ciao Corona study, a population-based study with randomly selected schools looking at seroprevalence of SARS-CoV-2 antibodies and clustering longitudinally in the canton of Zurich [8, 18, 19]. Invited schools were selected based on high incidence areas comprising the city of Zurich and one school of each of 4 adjacent districts to reach 600-800 children from primary and secondary schools. Herein, we use the term children for both primary and secondary school children and adolescents. Teachers could participate if they taught selected classes for at least 8 hours/week and a minimum of 2 days per week.

In each school, two testing days (T1 and T2) were done 1 week apart. Each time, two parallel buccal swabs were collected from children and teachers, 1 for the RDT (STANDARD Q® COVID-19 Ag Test from Biosensor/Roche) and 1 for the PCR (Analytica, Zurich, Switzerland). SARS-CoV-2 PCR was performed using the CE-IVD-marked Allplex™ SARS-CoV-2 Assay (Seegene Inc, Seoul, Republic of Korea) to detect the N-, S-, RdRP- and E-gene on a CFX96™ Real-time PCR System (Extraction-free method) [25] and was used as reference standard. PCR was done on buccal instead of a naso-pharyngeal swab. The agreement between PCR and RDTs using buccal swabs during acute SARS-CoV-2 infection has been validated in an adult symptomatic sample of 928 patients [26]. In this validation study, detection of acute infection was 39.8 and 40.1% for PCR on a buccal and naso-pharyngeal swab, respectively. Diagnostic performance of both PCRs was equivalent with reciprocal sensitivities of 95.7 and 96.5%. Sensitivities of the RDT compared to naso-pharyngeal PCR was 92.9% (86.4-96.9%), specificity was 100% (99.3-100%).

Children and teachers were instructed not to eat and drink for 1 hour prior to testing. The buccal swabs were taken in a large room where participants could be at least 2 m apart. Swabs were taken with closed mouth over 1 min. Two swabs were held closely together so that they were exposed to the same location of the enoral space. Swabbing of the mucosal membrane was done in the whole enoral space with some pressure application to make sure to have some cells contained in the swab. One swab was then used for the PCR and one for the RDT test. The RDT was performed directly in the schools and were read at 15-20 min and judged by two study team members in agreement as positive or negative. Each RDT biosensor was then photographed and the pictures saved on a secure university server. All positive RDT tests were repeated using a new buccal swab 2 hours to 2 days later (for four participants, also with the same buccal swab immediately when the signal was borderline positive to confirm). The two hours repeat was established after the first positive test experience. Acute symptoms during the last 5 days in children were assessed by staff members during testing prior to the swabs.

It was planned to inform parents and the school principals in case of positive RDTs in order to start TTIQ. After 3 false positive RDTs on the first day of testing, we changed procedures and required a confirmation by a repeated positive RDT and/or PCR test before initiating the isolation and quarantine measures by contact tracing teams.

The target sample size was calculated based on a number of assumptions: given that the population of 0-9 and 10-19 year-olds in the canton of Zürich is 161’613 and 141’482 respectively, weekly incidence of PCR diagnosed cases in 44^th^ week of 2020 was 80 in 0-9 year-old children and 692 in 10-19 year old adolescents, assuming that only 1 in 10 cases is diagnosed with PCR (as the ratio of tested children has seemingly increased substantially since spring 2020) [8], assuming that an asymptomatic infected child is PCR positive for 7 days, that 30% of undiagnosed cases are quarantined and do not attend school, we expected 1 PCR positive case in 308 children 0-9 years old and 1 in 32 children 10-19 year-old.

Therefore, in order to test the feasibility of the surveillance and maximize the probability of detecting positive cases, we oversampled secondary schools. 5 primary and 9 secondary schools were invited. Due to the character of the study, highly uncertain assumptions, the urge to proceed during the period of high incidence of SARS-CoV-2 infections, and the available resources and testing capacity, we aimed to enrol 700-1000 children in total. We invited all 1299 children from the already invited classes (regardless of previous participation in the seroprevalence study). With predicted participation rate similar to the other rounds of testing (50-60%), we expected 750-850 children in this study to take part.

## RESULTS

In total, 14 out of 15 invited primary and secondary schools agreed to participate. 641 from 1299 (49%) invited children from 67 invited classes took part at least one time (568 at T1, 602 at T2). Overall participation rate in at least one of the testing was 93% on the school level, 92% on class level, and 50% (interquartile range 35% to 65%, range 8% to 91%) on the child level within classes. Characteristics of study participants are given in Table 1. RDT and PCR were performed for all participating children and teachers. A repeat RDT and PCR in addition between week 1 and 2 was done in all initially positive children and teachers with a positive RDT.

**Table 1.** Characteristics of study participants

|  | Children | Teacher |
| --- | --- | --- |
| Number of participants | 641 | 66 |
| Sex, n (% female) | 370 (58) | 46 (70) |
| **Primary school** |  |  |
| Lower level, n (%) | 161 (25) | 29 (44) |
| Middle level, n (%) | 153 (24) |  |
| **Secondary school** |  |  |
| Upper level, n (%) | 327 (51) | 37 (56) |

None of the teachers and only 1 child had a positive PCR at any of the two testing visits, corresponding to a point-prevalence of 0.2% (CI 0% to 1.1%) at T1 and 0% at T2. This child with positive PCR at T1 had a cycle threshold value of 31-33 (N-Gen 31, RdRP-/S-Gen 32, E-Gen 33), was negative on the RDT at T1 and negative in PCR and RDT at T2. There were 7 (1.1%) positive RDTs in children and 2 (3.0%) positive RDT in teachers at T1 or T2 among 5 schools (overall point-prevalence 1.3%). 4 positive RDTs were also repeated immediately with the same buccal swab and remained positive. All 9 positive RDTs were also repeated 2 hours to 2 days later by taking a new buccal sample, and were negative both on RDT and PCR, respectively. The signals of the RDTs were in all cases weak-to-moderate (Figure 1), and were confirmed by two persons experienced in RDT testing and blinded to reported symptoms and our judgement.

**Figure 1:** Self-reported symptoms during the last 5 days in 198 of 567 (35%) children (1 child missing) at the initial T1 testing

198 (35%) children and 5 (8%) teachers reported mild symptoms during the 5 days prior to testing (Figure 2). Symptoms of week 1 (T1) and 2 (T2) did not differ significantly. The child with a positive PCR at T1 reported a runny nose at T1, but not on T2, when a loss of smell or taste was reported instead. 3 children, but no teachers reported mild symptoms.

**Figure 2:** The range of signal in the performed rapid diagnostic tests (STANDARD Q® COVID-19 Ag Test from Biosensor/Roche, Switzerland) showing the range of detected signals in our cohort of 641 children and 66 teachers with a moderate (A) and weak (B) signal of positive tests, as well as a negative test (C).

## DISCUSSION

In this nested study aiming to detect the spread of SARS-CoV-2 infections in schools at a time of high community transmission during the second wave of the pandemic (December 2020) in a country showing one of the highest incidences worldwide. Point-prevalence in children (0-0.2%) and teachers (0%) was low and clustering on the class or school level thus unlikely, given that participation of schools, classes and individual children and teachers in the testing was high. Given the very low point prevalence even in a setting of high incidence of SARS-CoV-2, the usefulness of a surveillance system for entire schools on an individual, regional or national level is questionable and currently not needed, especially with a well-functioning targeted TTIQ strategy in place for symptomatic children and school personnel. Specificity of the RDT in children was within the lower boundary of performance and needs further evaluation for its use as surveillance or targeted TTIQ strategy test in schools.

The very low point-prevalence of 0-0.2% of SARS-CoV2 infected children supports the debated political decision of many countries to leave schools open despite the striking numbers of confirmed cases during the second wave of the pandemic. Based on our power calculation, a high expected ratio of diagnosed to undetected cases (the SARS-CoV-2 seroprevalence of roughly 8% in children of the same schools from our main study in November 2020 with a dark figure of 1:8 [19], and the high case numbers in Switzerland at the peak of the second wave (i.e., 300 new symptomatic cases per 100’000 inhabitants between mid-November and mid-December 2020) we expected to detect at least a few asymptomatic or oligosymptomatic children or teachers in school, but we did not. Based on the relatively low number of tested children, one has to be careful in drawing too strong conclusions. Yet, this is a promising finding in consideration of the highly controversial and debated role of SARS-CoV-2 transmission in schools, [1-3] and given the fact that many experts support nation-wide school closures [2, 3].

Published modelling studies on the effect of school closures often rely on strong theoretical assumptions, can often not adequately control for important confounding because of their ecological nature, and although interesting from a scientific perspective, should not replace studies based on prospectively collected data. School closures should be the ultimate decision taken, as they lead to harmful global consequences on health, society and economics and dramatically increase existing inequalities [27, 28]. Before this is done, less dramatic decisions with schools remaining open can be taken, although current evidence for their effectiveness is not well studied [13]. It seems, that established Swiss procedures with school-based preventive measures (e.g., masks for teachers and children >12 years in the open space of schools, stable class constellations, tapering school breaks, the ban of group events such as school camps, physical distancing in class and teachers’ rooms), contact tracing and leaving febrile and/or highly symptomatic children at home, seem to work in hindering the SARS-CoV-2 spread by and through children in schools. So far, preliminary evidence suggests lower transmission from children to teachers than the other way around [29].

Our results suggest that surveillance of SARS-CoV-2 infections in schools is not efficient using PCR or RDTs. The prevalence of infections was very low under current protective measures and regulations of authorities so that many children and school personnel would need to be tested to identify single unrecognized cases. And even then it is unclear if such asymptomatic or oligosymptomatic children would infect others as transmission rates are low [29], and outbreaks infrequent and local [30-32]. Instead, acute virus testing of children seems more efficient only if they are at least moderately symptomatic [33]. These children should be sent home anyway but should be tested to trigger TTIQ.

There was one child with a positive PCR test in week 1 with a cycle threshold value of 31-33 closely above the detection threshold. The second test was negative in week 2, RDT tests were negative on both occasions. This picture suggests viral shedding during the postinfectious state [23], rather than a false negative RDT. If the child had had an infectious state during the first testing, in this case with a false negative RDT, one would expect to have at least a positive PCR test on week 2. This child was at school with just a runny nose. He/she was tested with an additional PCR two days after our first PCR due to a positive family member which was also positive. She/he then went into quarantine together with 3 classmates that had a close contact until our T2 testing. Interestingly, the child developed anosmia only at T2 when the PCR test had become negative. Although symptoms in this case were prolonged to 10 days at least, symptoms occurred during the post-infectious state. Despite the fact that this child was tested positive on PCR at T1 and went to school at least some days in an infectious state, there was no report of additional infections of classmates. It is possible that measures such as wearing masks, physical distancing and hygiene procedures were sufficient to prevent transmission.

Assuming PCR as gold standard reference method, the 9 false positive RDT tests were surprising although it is well known that they occur more frequently in low incidence settings like schools [34]. To differentiate between false and true positive is not always an easy task. The false positive result is highly likely in all 7 children and 2 teachers, as all repeat RDTs in a new buccal swab and likewise both PCRs collected in parallel to the RDTs were negative. The buccal instead of the naso-pharyngeal method has been validated in a large sample of adult outpatients with SARS-CoV-2 symptoms taking the naso-pharyngeal swab as gold-standard [26]. We cannot fully rule out false negative PCRs, as we did not measure human genetic material in the PCRs to confirm cellular content in the buccal swab. As the positive RDTs occurred in different schools at different time points in different age levels and were analyzed by different personnel that were all tested negative on RDTs taken at the beginning of the study, we could not detect any systematic factor that might have caused the false positive results. The negative RDTs in the repeats were promising. Taking a new buccal swab is important to minimize possible contamination. Our findings are important as surveillance and screening in the educational system in many countries nowadays extensively use RDTs without having a control PCR. The Centers for Disease Control and Prevention does not recommend screening testing in schools [35], mostly because real-world studies of its effectiveness are lacking. Yet, cost-effective and important and prompt decisions for children, families and schools could be taken for instance in children or teachers with unclear symptomatology or in outbreak situations, given that same-day results can prevent 80% of new transmissions, whereas a 7-day delay stops only 5% [36]. Specificity of RDT in our sample was 99.4% for children which is at the lower boundary of test performance reported by the company or validation studies in symptomatic adults [22] with reported specificity of 99.5% (95% CI 98.1% to 99.9%). The specificity was lower for the teachers, but due to the low number of tested teachers, one has to be very careful in judging test performance. Until RDTs can be used in surveillance or even with a targeted TTIQ strategy for children and school personnel, irrespective of symptoms, repeat testing in a new buccal swab and control PCR should be considered. So far, collection of saliva for SARS-CoV-2 diagnosis is increasingly recognized as valid alternative [37].

Strength of the study lies in the population-based sample of mostly asymptomatic children in schools tested in a country with one of the highest incidences of SARS-CoV-2 worldwide. Further strength of this study is the control of RDT by PCR using a prospective design. Limitations include the low prevalence of observed acute infections in the school system, which limited our capability to tease out the usefulness of RDTs as surveillance or a targeted TTIQ strategy test in schools.

## CONCLUSION

This is one of the few SARS-CoV-2 surveillance studies in asymptomatic or oligosymptomatic children in the school setting, documenting that unrecognized virus spread within schools is not a major problem even in a country with one of the highest SARS-CoV-2 incidence rates during the peak of the second wave in December 2020. Schools seem to be safe with the protective measures in place. A monitoring system to detect acute infections and possible clustering in schools even in a high incidence setting of Ciao Corona does not seem to be informative and thus not needed with the actual preventive measures in place.

## DATA AVAILABILITY STATEMENT

The raw data supporting the conclusions of this article will be made available by the authors, on reasonable request.

## Data Availability

NA

## AUTHOR CONTRIBUTIONS

MAP, SK, AU, TR and CB designed the study. SK and PA were responsible for recruitment of schools and communication with school principals. GPP, PA, SK and TR were responsible for testing at schools including supervision of testing personnel. SK coordinated the process of test interpretation of positive rapid diagnostic tests with external experts. AU carried out the statistical analysis. All authors contributed to the interpretation of study findings. SK wrote the first draft of the manuscript, and all authors critically reviewed and approved the final version of the manuscript.

## ACKNOWLEDGMENTS

Thanks to the contribution of participating children and schools we were able to run the study in such a short time. Not to forget the students and helpers that worked long hours. We thank Ruedi Jung for his support in data management, Nevesthika Muralitaharan for her administrative support, and Meret Fauchère for her coordination of the testing teams. We also thank the Cantonal and City authorities and the Cantonal Ethics Committee of Zurich for their responsiveness to our requests and their support.

